# Obesity paradox seen in cardiogenic shock with overweight and obesity having the lowest, whereas cachexia has the highest mortality using the large Nationwide Inpatient Sample (NIS) database

**DOI:** 10.1101/2025.05.05.25327024

**Authors:** Mohammad Reza Movahed, Mehrdad Mahalleh, Mehrtash Hashemzadeh

## Abstract

**Introduction:** The obesity paradox has been observed in patients with cardiovascular disease. The goal of this study was to evaluate if obesity has a protective effect in patients presenting with cardiogenic shock.

**Method:** Using a large Nationwide Inpatient sample (NIS) database, we evaluated mortality in patients with cardiogenic shock based on weight categories in adults.

**Results:** A total of 843,020 patients had a diagnosis of cardiogenic shock in the database over age 18. We found that overweight and obesity had the lowest mortality using univariate or multivariate analysis (overweight mortality of 20.66% vs obesity mortality of 26.6 % vs 34,3% of normal weights). In contrast, cachexia was associated with the highest mortality in univariate analysis (cachexia 40.4%). Using multivariate analysis adjusting for baseline characteristics and comorbidities, these relations remained unchanged (cachexia MVOR; 1.13, CI: 1.21-1.13, p <0.001, overweight MVOR: 0.52, CI; 0.43-0.65, p<0.001, obesity MVOR: 0.76, CI: 0.73-0.79, p<0.001). After multivariate adjustment, morbid obesity had similar mortality to patients with normal weight (morbid obesity MVOR: 0.99, CI 0.95-01.03, p=0.6)

**Conclusion:** We observe a partial obesity paradox in patients with cardiogenic shock, showing that overweight followed by obesity has the lowest mortality whereas cachexia has the highest mortality despite multivariate adjustment.

## Introduction

In most Westernized countries, including the United States, obesity has become a major public health concern, with three out of four Americans being overweight or obese [1–3]. Obesity negatively impacts cardiovascular health by increasing the risk of metabolic syndrome, diabetes, and dyslipidemia, particularly through elevated triglyceride levels. Obesity is associated with increased inflammation and, additionally, it raises the risk of coronary heart disease and hypertension, which potentially can lead to heart failure (HF). Furthermore, Obesity is linked to the occurrence of atrial fibrillation (AF), decreased renal function, increased risk of venous thromboembolism (VTE), and respiratory disease. These conditions, whether occurring separately or together, can harm the prognosis for HF [3, 4]. Despite the clear association between obesity and various cardiovascular diseases, current research has revealed the "obesity paradox.". This paradox postulates that individuals with higher BMIs have better outcomes and a lower death rate than those with normal BMIs [5]. In a meta-analysis involving individuals with acute coronary syndrome (ACS), Niedziela et al. [6] found that patients who were overweight, obese, or extremely obese had mortality rates that were considerably lower than those with normal body weights. Similar results have been observed in cases of septic shock and HF [7].

This paradox has been documented in both HF with reduced EF (HFrEF) and HFpEF, where overweight or mildly obese individuals exhibit decreased all-cause and cardiovascular mortality rates. However, hospitalizations tend to increase with severe obesity in this population [7, 8].

Conversely, in advanced HF, obesity may be associated with increased complications and lower survival, particularly in cases managed with heart transplantation or left ventricular assist devices [9, 10]. About 7%–10 % of individuals with acute myocardial infarction (AMI) or HF experience cardiogenic shock (CS), a severe and morbid complication [11, 12]. The existence of the obesity paradox in patients with CS is a topic of debate. Studies by Sreenivasan et al. [13] and Hashmi et al. [14] did not observe the obesity paradox in a significant US population with CS compared to non-obese individuals. These studies reported higher mortality rates in obese patients compared to non-obese individuals after CS. However, Patlolla et al. [15] and Chatterjee et al. [16], who employed a partially overlapped US database, observed that there was an obesity paradox associated with CS since they discovered noticeably decreased mortality in individuals with higher BMIs. Hermansen et al.’s study [17] in a Danish patient cohort revealed no relationship between obesity and the results of CS. In another study by Kwon et al. [18] in South Korea, it was found that men with obesity had a 37% lower death rate compared to non-obese men, whereas women did not show an association with obesity.

In CS, the relationship between weight category and obesity is not conclusively proven. Given the conflicting evidence and significant clinical implications, further investigation is needed to clarify the relationship between obesity and outcomes in CS. In this study, using the large Nationwide Inpatient Sample (NIS) database, we aimed to investigate whether obesity has a protective effect in patients presenting with CS.

## Methods

### Data Source

The Nationwide Inpatient Sample (NIS), the dataset included in this study, was created as part of the Healthcare Research and Quality (AHRQ) Project on Healthcare Cost and Utilization (HCUP). The NIS HCUP data is deidentified and publicly accessible, therefore, the study did not require approval from an institutional review board. Nearly a twenty percent stratified sample of discharges from community hospitals is included in the NIS. This study was exempt from IRB as it is publicly available without any patient identifiers.

### Study Population

Individuals who received their discharge from an NIS hospital between 2016 and 2020 and who were at least 18 years old were included in this retrospective research. International Classification of Diseases 10th Revision (ICD-10) codes were utilized to query and develop the study population using the NIS database.

### Study Outcomes

Mortality was the primary outcome of this study. In multivariate analysis, this outcome was adjusted for age, gender, race, and various variables including the history of smoking, chronic obstructive pulmonary disease (COPD), diabetes mellitus (DM), hypertension, chronic kidney disease (CKD), ST-elevation myocardial infarction (STEMI), non-ST-elevation myocardial infarction (NSTEMI), old Myocardial infarction (MI), and aortic valve surgery.

### Statistical Analysis

The demographic, clinical, and hospital characteristics of the patients are shown as 95% confidence intervals for categorical variables and as median (IQR) for continuous variables and proportions. To determine the odds of clinical outcomes over time as well as the odds of binary clinical outcomes with patient and hospital characteristics, logistic regression was used. For confounding, all statistical models were adjusted. All analyses were performed following the implementation of population discharge weights. Every p-value is two-sided, and a value of p<0.05 indicates statistical significance. STATA 17 (Stata Corporation, College Station, TX) was used for data analysis.

## Results

### Patient Characteristics

Table 1 provides a comprehensive overview of patient characteristics from 2016 to 2020, focusing on the total population of patients with CS and subsets categorized by weight: normal weight, cachexia, overweight, obesity, and morbid obesity.

**Table 1.**
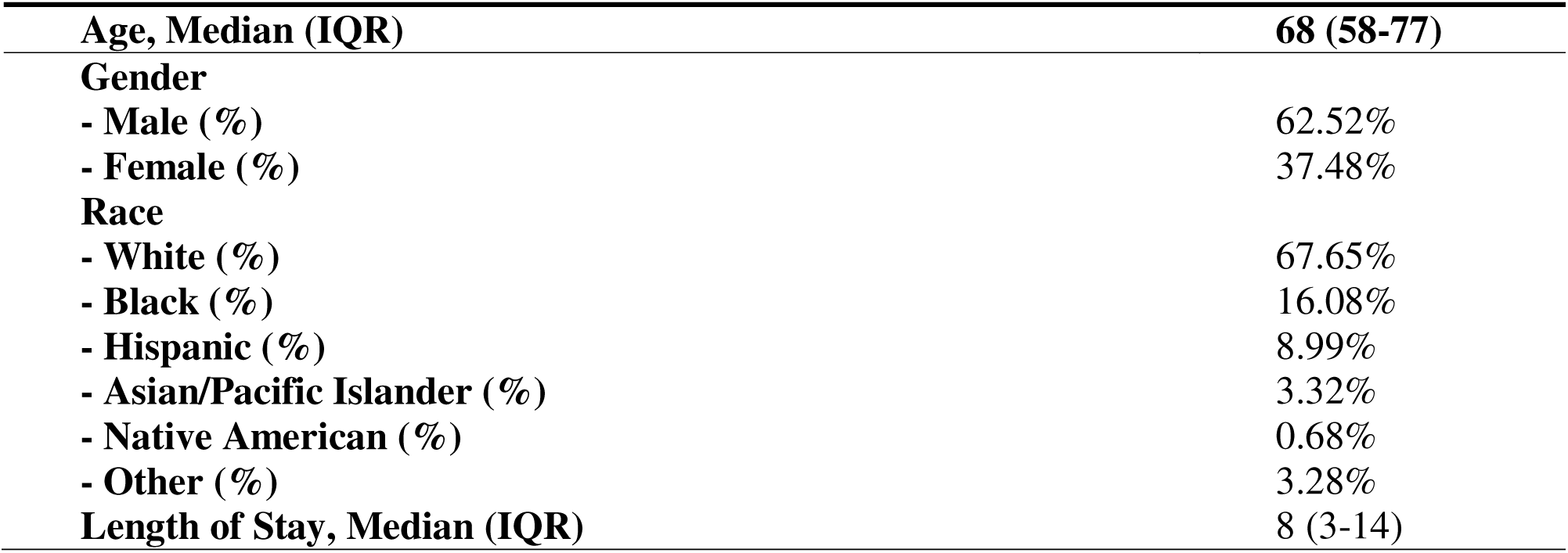
Baseline Characteristics of Patients with Cardiogenic Shock (n=843,020)

The total population comprised 843,020 patients, with 678,525 (80.5%) classified as having normal weight, 19,215 (2.3%) with cachexia, 3,175 (0.4%) who were overweight, 72,940 (8.7%) with obesity, and 70,150 (8.3%) with morbid obesity. The median age of the overall population was 68 years (IQR: 58–77). Among the subgroups, patients with cachexia had the highest median age at 70 years (IQR: 61–80), reflecting their advanced disease status, whereas patients with morbid obesity were the youngest, with a median age of 62 years (IQR: 53–70). In terms of clinical outcomes, the median length of stay (LOS) for the entire cohort was 8 days (IQR: 3–14). Patients with cachexia exhibited the longest median LOS at 9 days (IQR: 4–18), indicative of their more complex clinical needs, while overweight patients had the shortest LOS at 7 days (IQR: 4–7). Gender distribution revealed that the majority of the total CS population was male, accounting for 62.52% of cases. The racial composition showed that 67.65% of patients were White, followed by 16.08% Black, and 8.99% Hispanic. This detailed demographic and clinical profile in Table 1 provides valuable insights into the diverse characteristics and outcomes of patients with CS across different weight categories.

### Mortality

The mortality rates for patients with CS from 2016 to 2020 varied significantly across different weight categories when compared to normal-weight patients. The overall mortality rate for the CS population was 33.52%. As indicated in Table 2, the overall mortality rate for normal-weight patients was 34.38%. Patients with cachexia exhibited a markedly higher mortality rate of 40.44%, with a p-value of <0.001, indicating a statistically significant difference compared to normal-weight patients. The OR for mortality in cachexia patients was 1.30 (95% CI: 1.21-1.38). Conversely, overweight patients demonstrated a lower mortality rate of 20.66%. This difference was also statistically significant, with a p-value of <0.001. The odds ratio for mortality in overweight patients was 0.50 (95% CI: 0.40-0.61). Patients classified as obese had a mortality rate of 26.62%, significantly lower than that of normal-weight patients, with a p-value of <0.001. The OR for mortality in obese patients was 0.69 (95% CI: 0.66-0.72). Lastly, patients with morbid obesity had a mortality rate of 31.01%, which was significantly lower than the normal-weight group, with a p-value of <0.001. The OR for mortality in morbidly obese patients was 0.86 (95% CI: 0.83-0.89).

**Table 2.** Baseline Characteristics of Patients with Cardiogenic Shock by Weight Category.

|  | **Normal-weight**<br>**(n=678,525)** | **Cachexia**<br>**(n=19,215)** | **Overweight**<br>**(n=3,175)** | **Obesity**<br>**(n=72,940)** | **Morbid Obesity**<br>**(n=70,150)** |
| --- | --- | --- | --- | --- | --- |
| **Age, Median (IQR)** | 68 (59-78) | 70 (61-80) | 67 (57-75) | 65 (56-73) | 62 (53-70) |
| **Gender** |  |  |  |  |  |
| - % Male | 63.38% | 63.39% | 67.40% | 62.26% | 54.95% |
| - % Female | 36.72% | 36.61% | 32.60% | 37.74% | 45.05% |
| **Race** |  |  |  |  |  |
| - White (%) | 67.77% | 60.73% | 68.13% | 69.22% | 66.59% |
| - Black (%) | 15.51% | 23.25% | 12.85% | 15.60% | 20.45% |
| - Hispanic (%) | 9.03% | 7.47% | 13.01% | 9.62% | 8.20% |
| - Asian/Pacific Islander (%) | 3.61% | 4.30% | 2.60% | 2.06% | 1.53% |
| - Native American (%) | 0.69% | 0.65% | 0.00% | 0.61% | 0.71% |
| - Other (%) | 3.39% | 3.60% | 3.41% | 2.89% | 2.52% |
| **Length of Stay, Median (IQR)** | 8(3-14) | 9(4-18) | 7(4-7) | 8(4-14) | 9(4-16) |
| **Total Charges \$, Median** | 134,211 | 115,920 | 143,657 | 156,064.5 | 155,419 |

To ensure the robustness of the findings, the analysis of mortality rates for CS patients was conducted for each specific year from 2016 to 2020. The analyses revealed significant and consistent variations based on body weight categories. Cachexia patients consistently exhibited higher mortality rates compared to normal-weight patients throughout the years, with Ors ranging from 1.19 to 1.54 (Table 3). Conversely, overweight patients consistently showed significantly lower mortality rates than normal-weight patients across all years, with ORs ranging from 0.41 to 0.66 (Table 4). Similarly, obese patients demonstrated consistently lower mortality rates compared to normal-weight patients each year, with ORs ranging from 0.65 to 0.73 (Table 5). Morbidly obese patients also consistently exhibited lower mortality rates than normal-weight patients throughout the study period, with ORs ranging from 0.80 to 0.90 (Table 6). In the multivariate analysis (Table 7), adjusting for baseline characteristics and comorbidities from 2016 to 2020, the impact of weight categories on mortality revealed distinct patterns. Cachexia remained associated with a higher mortality rate, with an OR of 1.21 (95% CI: 1.13– 1.30, p < 0.001), indicating a significantly increased risk compared to normal-weight individuals. Overweight and obesity showed reduced mortality risks, with ORs of 0.52 (95% CI: 0.43–0.65, p < 0.001) and 0.76 (95% CI: 0.73–0.79, p < 0.001), respectively. However, the association for morbid obesity was no longer statistically significant after adjustment, with an OR of 0.99 (95% CI: 0.95–1.03, p = 0.683). These adjusted results suggest that the increased mortality risk associated with cachexia and the reduced risk linked to overweight and obesity are robust, while the effect of morbid obesity on mortality does not show a significant difference compared to normal weight after accounting for other variables.

**Table 3.** Mortality Rates and Odds Ratios by Weight Category in Patients with Cardiogenic Shock.

| **Weight Category** | **Mortality Rate (%)** | **Univariate OR (95% CI)** | **Multivariate OR (95% CI)** | **P-Value (Univariate)** | **P-Value (Multivariate)** |
| --- | --- | --- | --- | --- | --- |
| **Normal Weight** | 34.38 | Reference | Reference | - | - |
| **Cachexia** | 40.44 | 1.30 (1.21-1.38) | 1.35 (1.25-1.45) | <0.001 | <0.001 |
| **Overweight** | 20.66 | 0.50 (0.40-0.61) | 0.55 (0.44-0.66) | <0.001 | <0.001 |
| **Obesity** | 26.62 | 0.69 (0.66-0.72) | 0.72 (0.69-0.75) | <0.001 | <0.001 |
| **Morbid Obesity** | 31.01 | 0.86 (0.83-0.89) | 0.90 (0.87-0.93) | <0.001 | <0.001 |

**Table 4.** Annual Mortality and Odds Ratios in Cardiogenic Shock: Cachexia vs. Normal Weight (2016-2020)

| **Year** | **Mortality**<br>**(Cachexia)** | **Mortality (Normal**<br>**Weight)** | **P-**<br>**value** | **Odds Ratio**<br>**(C.I)** |
| --- | --- | --- | --- | --- |
| **2016** | 46.64% | 36.21% | <0.001 | 1.54 (1.28-1.85) |
| **2017** | 41.52% | 35.72% | 0.004 | 1.27 (1.08-1.50) |
| **2018** | 37.95% | 34.01% | 0.02 | 1.19 (1.02-1.37) |
| **2019** | 39.84% | 32.46% | <0.001 | 1.38 (1.19-1.59) |
| **2020** | 38.96% | 34.21% | 0.001 | 1.23 (1.08-1.39) |

**Table 5.** Annual Mortality and Odds Ratios in Cardiogenic Shock: Overweight vs. Normal Weight (2016-2020)

| **Year** | **Mortality (Overweight)** | **Mortality (Normal Weight)** | **P-value** | **Odds Ratio (C.I)** |
| --- | --- | --- | --- | --- |
| **2016** | 20.59% | 36.21% | 0.009 | 0.46 (0.25-0.82) |
| **2017** | 20.91% | 35.79% | 0.001 | 0.47 (0.30-0.75) |
| **2018** | 17.39% | 34.01% | <0.001 | 0.41 (0.27-0.62) |
| **2019** | 18.11% | 32.46% | <0.001 | 0.46 (0.30-0.70) |
| **2020** | 25.60% | 34.21% | 0.05 | 0.66 (0.44-0.99) |

**Table 6.** Annual Mortality and Odds Ratios in Cardiogenic Shock: Obesity vs. Normal Weight (2016-2020)

| **Year** | **Mortality (Obesity)** | **Mortality (Normal Weight)** | **P-value** | **Odds Ratio (C.I)** |
| --- | --- | --- | --- | --- |
| **2016** | 27.44% | 36.21% | <0.001 | 0.67 (0.60-0.74) |
| **2017** | 26.62% | 35.79% | <0.001 | 0.65 (0.59-0.72) |
| **2018** | 26.58% | 34.01% | <0.001 | 0.70 (0.64-0.77) |
| **2019** | 25.25% | 32.46% | <0.001 | 0.70 (0.64-0.77) |
| **2020** | 27.38% | 34.21% | <0.001 | 0.73 (0.67-0.79) |

**Table 7.**
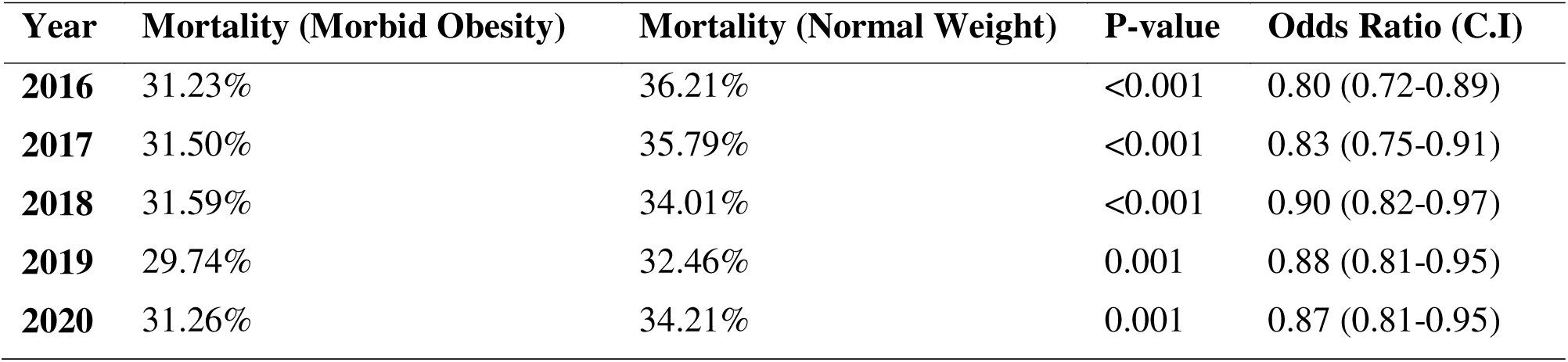
Annual Mortality and Odds Ratios in Cardiogenic Shock: Morbid Obesity vs. Normal Weight (2016-2020)

| **Year** | **Mortality (Morbid Obesity)** | **Mortality (Normal Weight)** | **P-value** | **Odds Ratio (C.I)** |
| --- | --- | --- | --- | --- |
| **2016** | 31.23% | 36.21% | <0.001 | 0.80 (0.72-0.89) |
| **2017** | 31.50% | 35.79% | <0.001 | 0.83 (0.75-0.91) |
| **2018** | 31.59% | 34.01% | <0.001 | 0.90 (0.82-0.97) |
| **2019** | 29.74% | 32.46% | 0.001 | 0.88 (0.81-0.95) |
| **2020** | 31.26% | 34.21% | 0.001 | 0.87 (0.81-0.95) |

## Discussion

The present study aimed to elucidate the association between weight categories and mortality in patients with CS using a large, nationally representative dataset. Our findings contribute to the ongoing debate regarding the obesity paradox in CS by providing a comprehensive analysis of mortality rates across different weight categories. Patients with cachexia had the highest mortality rates, significantly exceeding those of normal-weight patients. Conversely, overweight and obese patients demonstrated significantly lower mortality rates compared to their normal-weight counterparts. Our study supports the existence of an obesity paradox similar to that observed in coronary artery disease (CAD), ACS, and HF [19–22], albeit with a more complex relationship between weight categories and mortality than previously reported.

Prior research has yielded inconclusive findings regarding the relationship between obesity and outcomes in CS. A study analyzing data from the National Readmission Database did not observe an obesity paradox and showed that patients with class II (BMI 35.0–39.9 kg/m²) and class III obesity (BMI ≥40.0 kg/m²) had a greater risk of in-hospital mortality compared to those who were not obese (BMI 20.0–29.9 kg/m²) [13]. In contrast, in another study focusing on CS secondary to acute myocardial infarction (AMI-CS) admissions, which included about 340,000 cases, the in-hospital mortality rate was found to be lower for both underweight and overweight/obese individuals [15]. These discrepancies were further reinforced by another study on 54,044 patients with AMI-CS, which revealed that while obesity was associated with a lower risk of in-hospital mortality, severe obesity was linked to a higher risk compared to non-obese individuals [23]. Another prior study [24] conducted on a large group of patients with CS discovered a U-shaped relationship between BMI and in-hospital mortality, which did not confirm the obesity paradox in CS. The study revealed that the lowest mortality rates were recorded among individuals with BMIs within the middle range. The discrepancies between our findings and those of earlier studies may lie in variations in study populations and methods. This highlights the need for further research to clarify how truly weight categories influence mortality in diverse patient populations.

Adipose tissue has a significant function in producing different hormones and cytokines, such as adiponectin, which has a protective impact on the cardiovascular system against inflammation and systemic atherosclerosis [25, 26]. This may help explain the fact that obesity offers cardioprotective benefits in certain circumstances. While the weight category may not directly correlate with body fat composition or adipose tissue function [27], it might potentially be associated with the cardioprotective mechanisms observed in obese individuals. However, our study also found that while morbidly obese patients initially showed lower mortality rates, this association was no longer significant after adjusting for baseline characteristics and comorbidities. This suggests that the apparent survival benefit in morbidly obese patients might be confounded by other factors such as age, gender, and comorbid conditions. The lack of a protective effect in morbidly obese patients after adjustment indicates that extreme obesity could negate the survival advantages seen in moderately obese individuals, possibly due to the higher burden of comorbidities and the increased challenges in managing these patients.

The findings of this study have several implications for clinical practice. Firstly, they highlight the need for a different approach to the management of obese patients with CS. While moderate obesity appears to offer a survival benefit, extreme obesity does not. This issue shows the importance of individualized care strategies that consider the entire clinical profile of the patient. Clinicians should be aware of the potential benefits of moderate obesity in the context of CS but also remain careful about the increased risks associated with morbid obesity. Moreover, the high mortality rates in cachectic patients emphasize the need for early identification and intervention in this vulnerable population. Strategies aimed at nutritional support and muscle preservation could potentially improve outcomes in cachectic patients with CS.

## Limitations

This study has several limitations. This study is a retrospective observational study, which means it cannot establish cause-and-effect correlations. It is important to note that there may still be unmeasured factors that might influence the results, known as residual or unmeasured confounding. In addition, the retrospective nature of the analysis and reliance on administrative data may introduce selection bias and misclassification errors. The use of BMI as the sole measure of obesity does not account for variations in body composition, such as muscle mass versus fat mass, which could influence outcomes. Additionally, the dataset lacks detailed information on the severity of comorbidities and the specific treatments received, which could affect mortality rates.

Future research should focus on prospective studies that incorporate more precise measures of body composition and investigate the underlying mechanisms driving the obesity paradox in CS. Further studies are also needed to explore the impact of weight management interventions on outcomes in this population. Understanding the pathophysiological differences between moderate and extreme obesity could help tailor therapeutic strategies and improve prognostic assessments.

## Conclusion

In summary, our study provides robust evidence supporting the partial obesity paradox in patients with CS, with overweight and moderately obese patients exhibiting lower mortality rates compared to those of normal weight. However, this protective effect diminishes in the context of morbid obesity. These findings indicate the complexity of the relationship between obesity and mortality outcomes and highlight the importance of personalized care strategies in managing patients with CS. Addressing the high mortality in cachectic patients remains a critical area for future research and clinical intervention.

## Data Availability

it is nis data base pulicaly available

